# Multiclass Semantic Segmentation of Immunostained Breast Cancer Tissue with a Deep-Learning Approach

**DOI:** 10.1101/2022.08.17.22278889

**Authors:** Mauricio Alberto Ortega-Ruiz, Edgar Roman-Rangel, Constantino Carlos Reyes-Aldasoro

**Affiliations:** giCentre, Department of Computer Science, School of Mathematics, Computer Science and Engineering, City, University of London, UK; Universidad del Valle de Mexico, UVM; Computer Science Department. Instituto Tecnologico Autonomo de Mexico, ITAM. Mexico City

**Author notes:** {, }. {.

**Keywords:** Breast Cancer, Semantic Segmentation, Convolutional Neural Networks, BCSS Challenge

## Abstract

This paper describes a multiclass semantic segmentation of breast cancer images into the following classes: Tumour, Stroma, Inflammatory, Necrosis and Other. The images were stained with Haematoxilin and Eosin and acquired from the Cancer Genome Atlas through the Breast Cancer Semantic Segmentation Challenge. Over 12,000 patches of data and classes were generated from training images with the use of data augmentation.The segmentation was obtained with a U-Net architecture for which the hyperparameters were explored systematically. Optimal values were obtained with batch size = 8, Loss function Adam and 50 epochs, which took over 50 hours to train. Due to this fact and limitations in time, the rest of the parameters were explored with 10 epochs and we speculate that values would increase if 50 epochs would be used. The trained U-Net was applied to unseen images, per-patch and the following metrics were obtained from full scale WSI; Accuracy, Mean Area Under the Curve and Dice Index. No post-processing was applied. The resulting segmentations outperformed the baseline in terms of accuracy for some tissues; Tumour from 0.804 to 0.913, Inflammatory from 0.743 to 0.8364. The data is available from the Grand Challenges website (*https://bcsegmentation.grand-challenge.org/*) and the code is available from the following GitHub repository (*https://github.com/mauOrtRuiz/Breast_Cancer_Sem_Seg*).

## 1 Introduction

Breast cancer is one of the most common diseases worldwide and has a high mortality rate. An early breast cancer diagnosis is fundamental for improving the patients’ outcome [17]. Whilst new techniques like Radiomics and Liquid biopsies are promising, biopsies and histopathological examination remain the main tool to confirm the diagnosis of breast cancer [19]. Recently, computational and algorithmic approaches, under the term *Computational Pathology* have emerged as a strong tool to help pathologists in the diagnosis, prognosis and other tasks [10]. This area of research has grown significantly and has contributed widely from tasks like counting nuclei to outcome prognosis [7, 9, 14, 11]. There are many techniques employed to stain tissue, and perhaps the most common of all is the process called Haematoxylin and Eosin (H&E). The output of H&E will have stained nuclei in blue/purple (the *H*) and the cytoplasm in pink (the *E*) [3]. There exists a large number of different staining techniques, which exploit the use antibodies bind in different sites of the tissue and thus are more specific and can provide different information to that of H&E. Examples of these are CD31 [21], Ki67[13], CD133 [18], Masson’s Trichrome [2] in the tissue are more specific and can complement the use of H&E [4]. The advances in computational pathology are promising and these may soon impact directly in the diagnosis and treatments of many diseases [15].

Image segmentation of histopathology images is a key task and widely used for tumour detection [16]. Nuclei and cell Segmentation of histopathology images are basic steps required in histopathology analysis, as well as feature extraction and classification [6, 12, 26]. This full process gives support physicians to diagnosis cancer, quantify tumour size, cancer grading and even for treatment planning [16]. Segmentation has been studied based on traditional computer vision, machine learning and deep learning techniques. Traditional methods are mainly based on edge and region analysis, for example region growing and thresholding. In machine learning features are extracted based on computer image analysis, and data processing methods are next used to classify tissue images. Deep learning methods have reported outstanding results and are mainly based on different neural networks which need to be trained to determine certain network output (classification or segmentation). U-Net is a network frequently used for segmentation task [16]. However its performance depends on the variance of the several visual patterns that might be present in the images, and the amount of images available for robust learning of those patterns during a training process. In segmentation, reference data need to be prepared and hand delimited only by pathology specialists, which is a tedious process that might be affected by human errors. Due to the lack of segmentation training data different approaches have been proposed. Synthetic generation data is presented by [8] to automatically generates reference data. An automated weakly label assignment is presented by [28] and a teacher-student model for delineation of cancer region is given by [27]. Also, different datasets presented during challenge contests also address this task. For example, PAIP 2019 hepatocellular carcinoma segmentation and CAMELYION 2016 for metastasis detection in lymph nodes. Other segmentation regions have been also studied, like tumour segmentation [20, 22, 5], epithelial, stromal tissue discrimination [24], and cytoplasm [25]. Breast cancer Semantic segmentation challenge presents breast cancer data label images classified in 22 different regions by a set of pathology specialists [1], this data is used in present paper.

This paper presents a segmentation method for breast cancer images based on the U-Net architecture [23] with parameters carefully adjusted for optimal segmentation performance. Segmentation is satisfactory applied on 4 different regions: tumour, stroma, inflammatory, and necrosis. Results presented in this work outperforms two tissue regions presented in [1]. An adequate hyper-parameters analysis and tuning indicate that higher segmentation accuracy (0.9130) before can be achieved. Also, a higher number of training images contributed to this performance.

The remaining of this paper is organised as follows. Section 2 presents the description of the dataset obtained from BCSS Challenge, as well as the segmentation model used in this work. Section 3 presents the results obtained after evaluating different parameters of the model. Finally, we conclude in section 4.

## 2 Materials and Methods

### 2.1 Materials

The materials used in this paper consisted of whole-slide images (WSI) from the Breast Cancer Semantic Segmentation (BCSS) Challenge, which is available through the Grand Challenges website (*https://bcsegmentation.grand-challenge.org/*).

The dataset consists of 151 histologically-confirmed breast cancer WSI that were stained with haematoxylin and eosin (H&E). The images had different sizes and resolution. The majority (138 WSI) were acquired at a magnification of 40*×* and only 13 WSI at magnification of 20 *×*. These slides are part of the The Cancer Genome Atlas Program and were manually annotated by pathologists, pathology residents, and medical students through a crowdsourcing process [1]. The annotation was used to produce more than 20, 000 segmentation annotations in 22 different tissue regions.

For this work, only 4 tissue regions were selected: *Tumour, Stroma, Inflammatory*, and *Necrosis*, all other cells of the tissue are grouped into a category called *Other*. We use a 3D tensor structure (a cube) to represent each ground-truth, i.e., the expected output of a segmented image. This structure has the same size as its corresponding input image, and consists of five channels in depth, which mark the presence of a pathology in a given pixel, i.e., and indicator function. A representative sample of the data with one WSI, a Region of Interest (ROI) and annotations done by the specialists are illustrated in Fig. 1. From the full set of WSI, 15 images were randomly selected and used as test set. After resizing each image by a factor of 1*/*4, small patches of 256 *×* 256 pixels were extracted allowing overlap of 50%, i.e., 126 pixels. Samples of these patches are illustrated in Fig. 1. After data augmentation based on rotating 90 and 180 degrees, flip-up and flip-left, 12, 930 patches were obtained to train the U-Net. from which, 10,736 were used as training, 2,194 as validation, and 15 WSI were used as the test set. Table 1 summarises the materials used in this work.

**Fig. 1.**
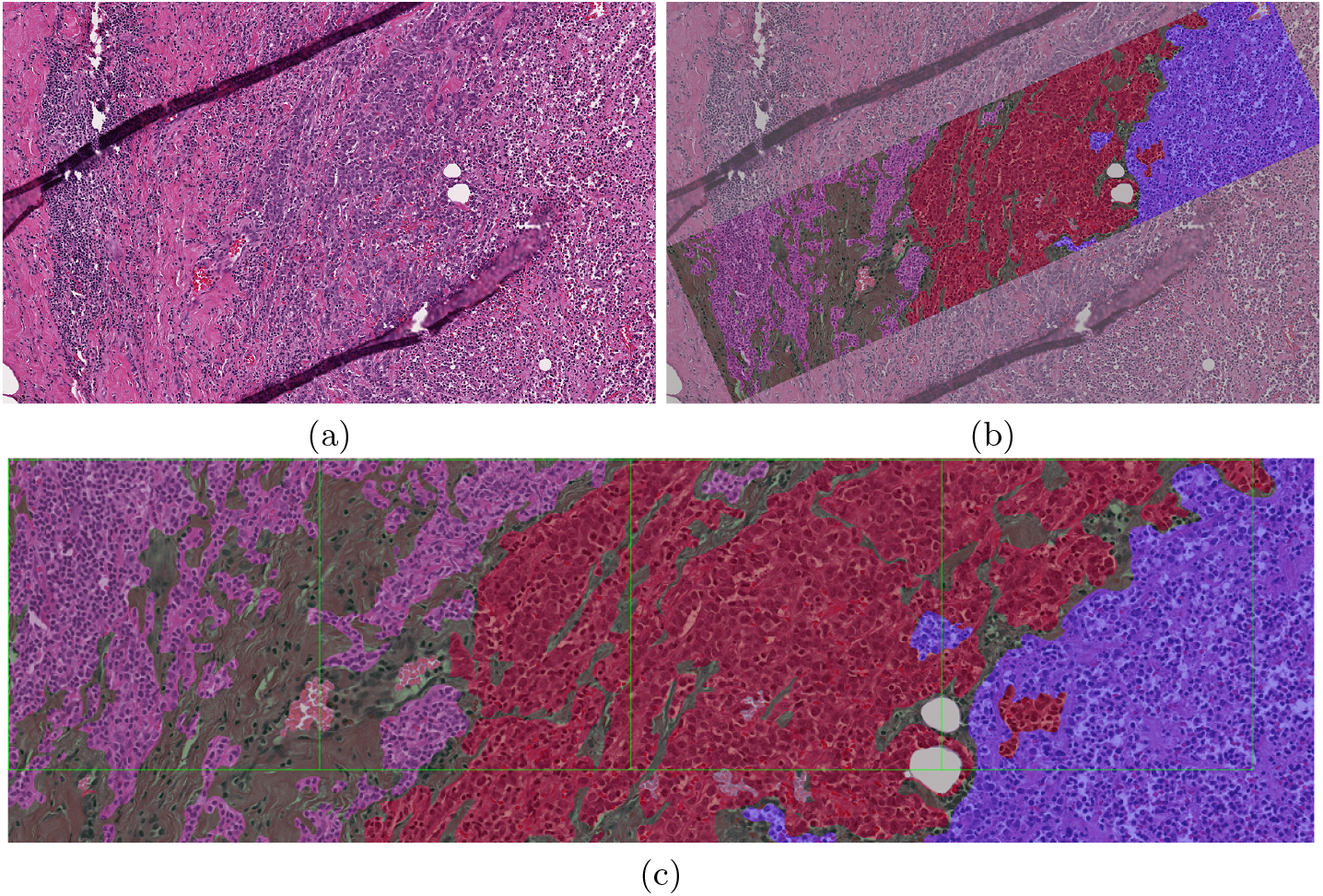
(a) One representative sample H&E image taken from BCSS challenge dataset. (b) A rectangular region of interest region annotated by specialists. The tissue regions selected in this work are tumour (red), stroma (green), inflammatory (purple), necrosis (blue), and other (gray). (c) Magnification of the region of interest with lines that indicate patches that will be extracted. It should be noted that tumour region is easier to distinguish, whereas stroma and necrosis are irregular regions and harder to distinguish.

**Table 1.**
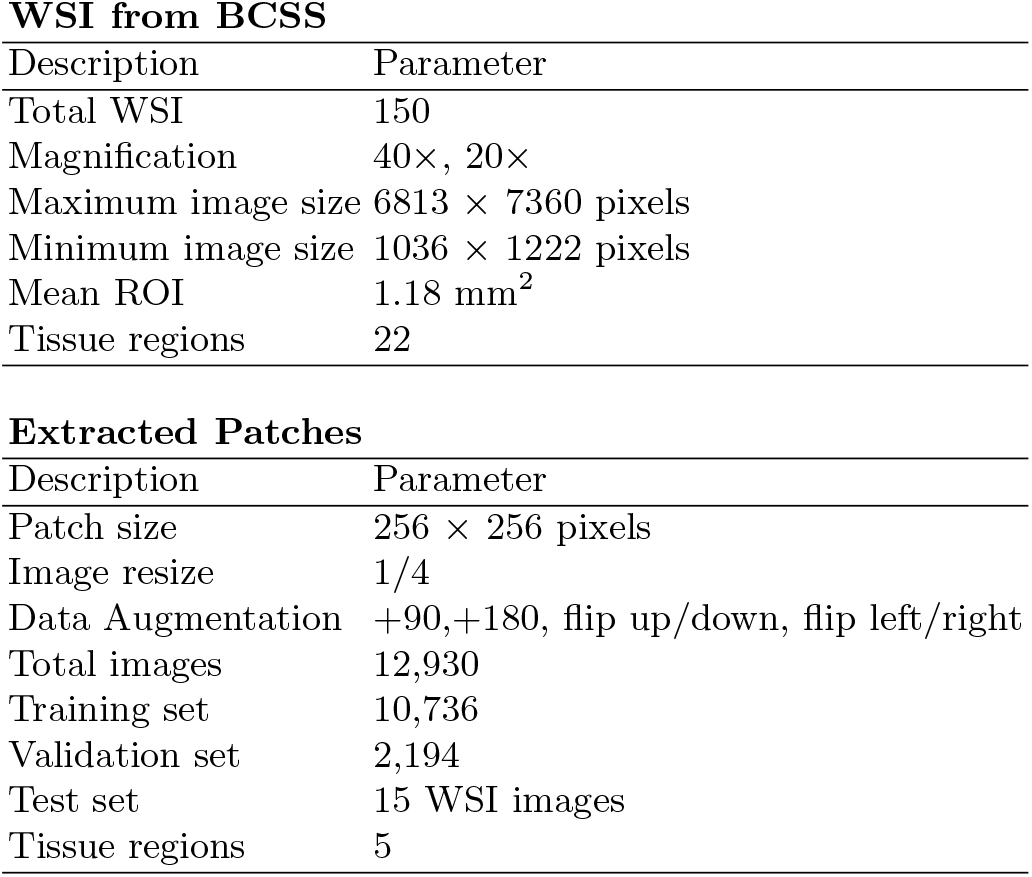
Characteristics of the dataset used in this work, first lines indicate original Whole Slide Images (WSI) parameters provided by Breast Cancer Semantic Segmentation (BCSS) and lower parameters describe patches used in this work.

**WSI from BCSS**
| Description | Parameter |
| --- | --- |
| Total WSI | 150 |
| Magnification | 40×, 20× |
| Maximum image size | 6813 × 7360 pixels |
| Minimum image size | 1036 × 1222 pixels |
| Mean ROI | 1.18 mm<sup>2</sup> |
| Tissue regions | 22 |

| Description | Parameter |
| --- | --- |
| Patch size | 256 × 256 pixels |
| Image resize | 1/4 |
| Data Augmentation | +90,+180, flip up/down, flip left/right |
| Total images | 12,930 |
| Training set | 10,736 |
| Validation set | 2,194 |
| Test set | 15 WSI images |
| Tissue regions | 5 |

### 2.2 Methods

A semantic segmentation of the WSI was obtained with a U-Net architecture [23]. The well-known U-Net architecture or simply *U-Net* is a Convolutional Neural Network (CNN) that has proven to be an excellent tool for semantic segmentation, which is approached phrasing it as a pixel-wise classification problem.

U-Nets consist of encoder-decoder sections. They contain a series of down-sampling steps (encoder) obtained by convolutions and downsampling operations, and then upsampling steps (decoder) formed by upsampling plus convolution operations. The most common formulation of this architecture contains as many upsampling steps as there are downsampling steps, such that the decoder can be understood as a reflected formulation of the encoder, and thus giving the name U-Net. Moreover, there are residual connection between pairs of convolutional layers, directly connecting the encoder with the decoder at the same depth within the model [23]. Fig. 3 shows a diagram of the U-Net model used in this work.

**Fig. 2.**
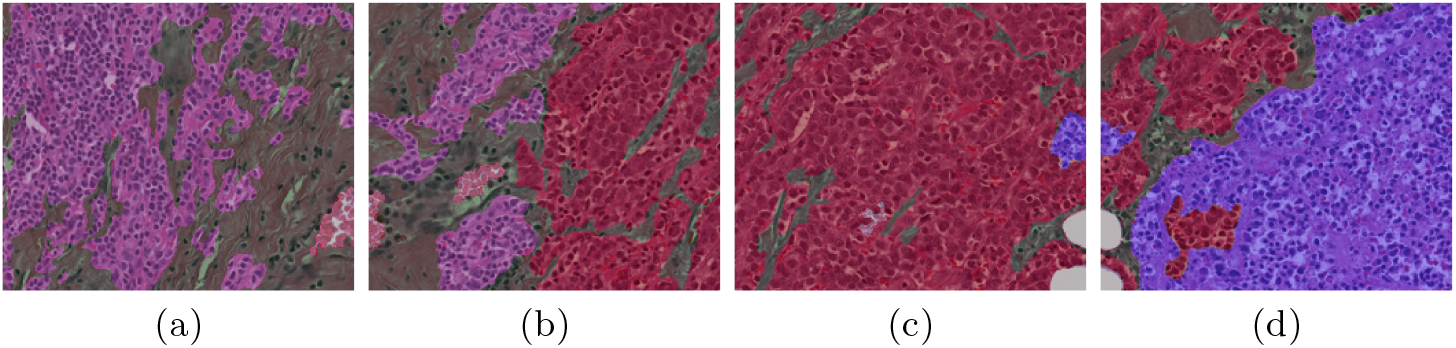
Four representative images extracted from image above 1. After applying data augmentation patches are used to train the neural network. Image shows separated patches, but also patches overlapped 126 pixels were extracted.

**Fig. 3.**
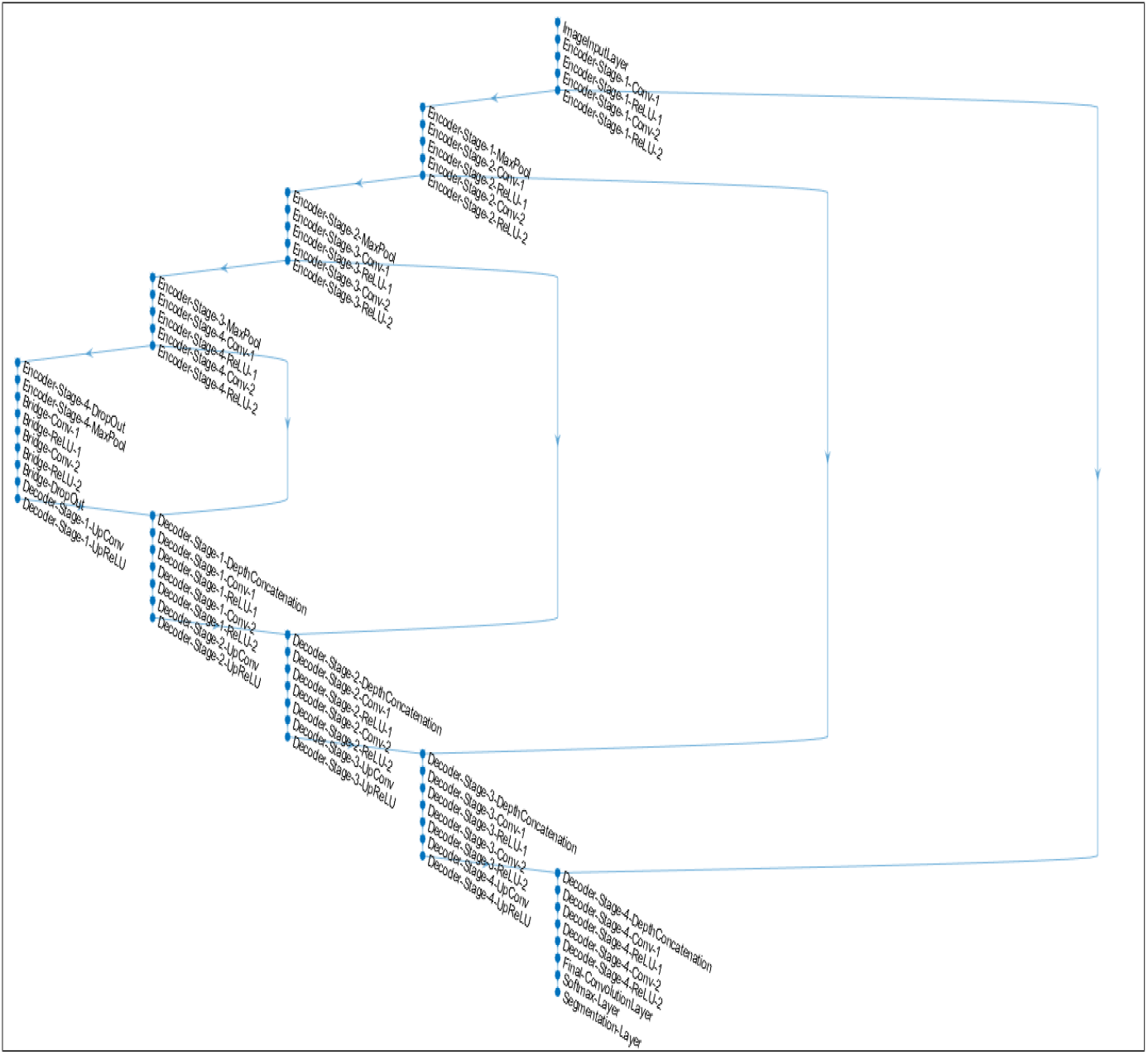
Illustration of the U-Net architecture used for the semantic segmentation of the breast cancer histological slides.

We use a U-Net network with depth 4, i.e., four layers in the encoder and four in the decoder, for performing semantic segmentation of the image set. All convolutional layers have 2D filters of size 3*×* 3 with ReLU activation functions, and they are followed by either max-pooling or upsampling layers with stride 2. The last layer consists of five convolutional filters, one for each class considered in this work, inclusing the class *other*. These filters are also of size 3 *×* 3, and they use a sigmoid activation function.

Different training parameters were systematically explored to optimise accuracy performance in both the training and validations sets, i.e., avoiding overfitting. Mini-batch size during training iterations was tested from 1 to 16 and the best results were provided by a mini-batch of size 8. Three loss functions were evaluated: stochastic gradient descent with momentum (sgdm), root mean square propagation (rmsprop), and adaptive moment estimation (adam). Adam provided the best segmentation results.

Also, training scenarios of 5, 10, 30 and 50 epochs were tested. Whilst the best results were provided by 50 epochs, this case took over 50 hours training in a Computer with processor Intel core i7-7700k, 16 GB RAM and CPU at 4.20 GHz with GPU. Thus, to explore all other parameters 10 epochs were selected.

### 2.3 Calculation of Accuracy, Dice coefficient and Area Under the Curve

The accuracy of the semantic segmentation was calculated on a pixel-based level. Pixel whose class was correctly predicted were counted as Correct, which included True Positive (*TP*) and True Negative (*TN*). Otherwise, incorrect pixels included False Positive (*FP*) and False Negative (*FN*). Therefore the **accuracy** was calculated as (*TP* + *TN*)*/*(*TP* + *TN* + *FP* + *FN*). **Dice coefficient** is calculated as (2*TP*)*/*(2*TP* + +*FP* + *FN*). Finally, the Receiver operating characteristic (ROC) curve followed the true positive rate (*TPR* = (*TP*)*/*(*TP* + *FN*)) against the false positive rate (*FPR* = (*TN*)*/*(*TN* + *FP*)) from which the **area under the curve** was calculated.

## 3 Results and Discussion

A summary of the exploration of the parameters is presented in Table 2. The accuracy is presented as the average across all five classes.

**Table 2.** Parameters evaluated during network training. A new U-Net training was done for every single parameter and accuracy was measured for the validation set after averaging on the overall tissue regions

| Batch Size |  |  |  |  |  |
| --- | --- | --- | --- | --- | --- |
|  | 1 | 2 | 4 | 8 | 16 |
| Validation | 0.5240 | 0.5275 | 0.5376 | 0.6900 | 0.6717 |
| Training | 0.4038 | 0.4100 | 0.7424 | 0.7277 | 0.7053 |
| Loss function |  |  |  |  |  |
|  | sgdm |  | adam |  | rmsprop |
| Validation | 0.6900 |  | 0.7610 |  | 0.7470 |
| Training | 0.7277 |  | 0.8745 |  | 0.8534 |
| epochs |  |  |  |  |  |
|  | 5 | 10 | 30 | 50 |  |
| Validation | 0.6321 | 0.6900 | 0.7640 | 0.9040 |  |
| Training | 0.8196 | 0.8745 | 0.8860 | 0.9571 |  |

**Table 3.** Comparison of the Baseline results [1] and the results obtained in this work with those values where the methodology proposed here outperformed the baseline are highlighted in **bold**.

|  | Baseline |  |  | This work |  |  |
| --- | --- | --- | --- | --- | --- | --- |
|  | Mean AUC | Dice | Accuracy | Mean AUC | Dice | Accuracy |
| Overall | 0.945 | 0.888 | 0.799 | 0.8401 | 0.7852 | 0.7395 |
| Tumour | 0.941 | 0.851 | 0.804 | 0.9129 | **0.8814** | **0.9130** |
| Stroma | 0.881 | 0.800 | 0.824 | 0.8395 | **0.8107** | 0.8043 |
| Inflammatory | 0.917 | 0.712 | 0.743 | 0.8882 | **0.8073** | **0.8364** |
| Necrosis | 0.864 | 0.723 | 0.872 | 0.8361 | 0.8320 | 0.7296 |
| Other | 0.885 | 0.666 | 0.670 | 0.7235 | 0.5945 | 0.4140 |

The parameters that provided the best results were: minibatch = 8, epochs = 50, and optimiser = adam. Namely, we observed that the performance increased with respect to the batch size up to 8 instances per batch, and with respect to the number of training epochs. Furthermore, the efficient approximation of the mean and variance of the gradients, that Adam implements, provides better results with respect to the less smoothed approximations of SGDM and RMSProp.

The network trained with the parameters previously mentioned was used to segment the remaining 15 WSI reserved as unseen test data. For this process, the images were resized by how 1*/*4 and divided in 256*×*256 sections to perform segmentation and were subsequently reassembled to the original resolution. No overlapping or post-processing was applied. Two cases of this process are presented in Fig. 5 and Fig. 6. In both cases Fig. (a) is the original H&E WSI, Fig. (b) is the Ground truth provided by the BCSS and Fig. (c) is the segmented imaged obtained by the method presented. Concordance between GT and segmented result are shown in white pixels in Fig. (d). Accuracy in Fig. 5 is 0.8297 and in Fig. 6 of 0.9509.

**Fig. 4.**
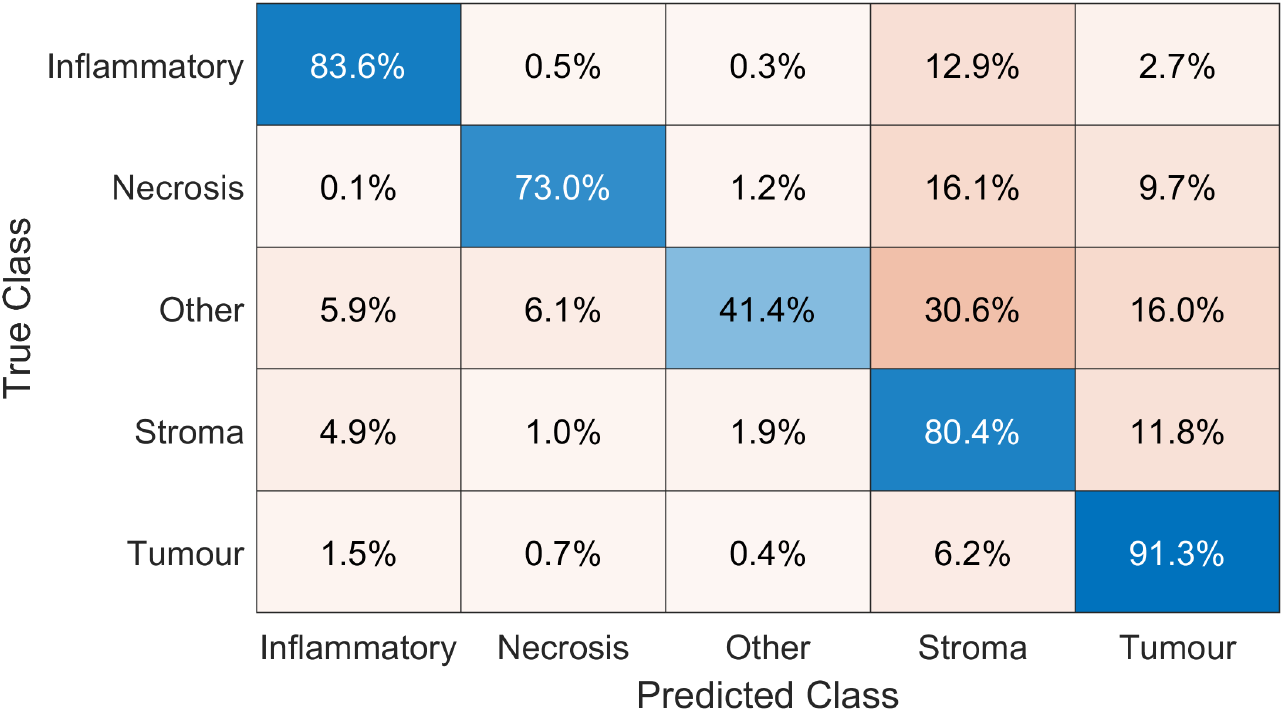
Confusion Matrix for the five classes into which the images were segmented.

**Fig. 5.**
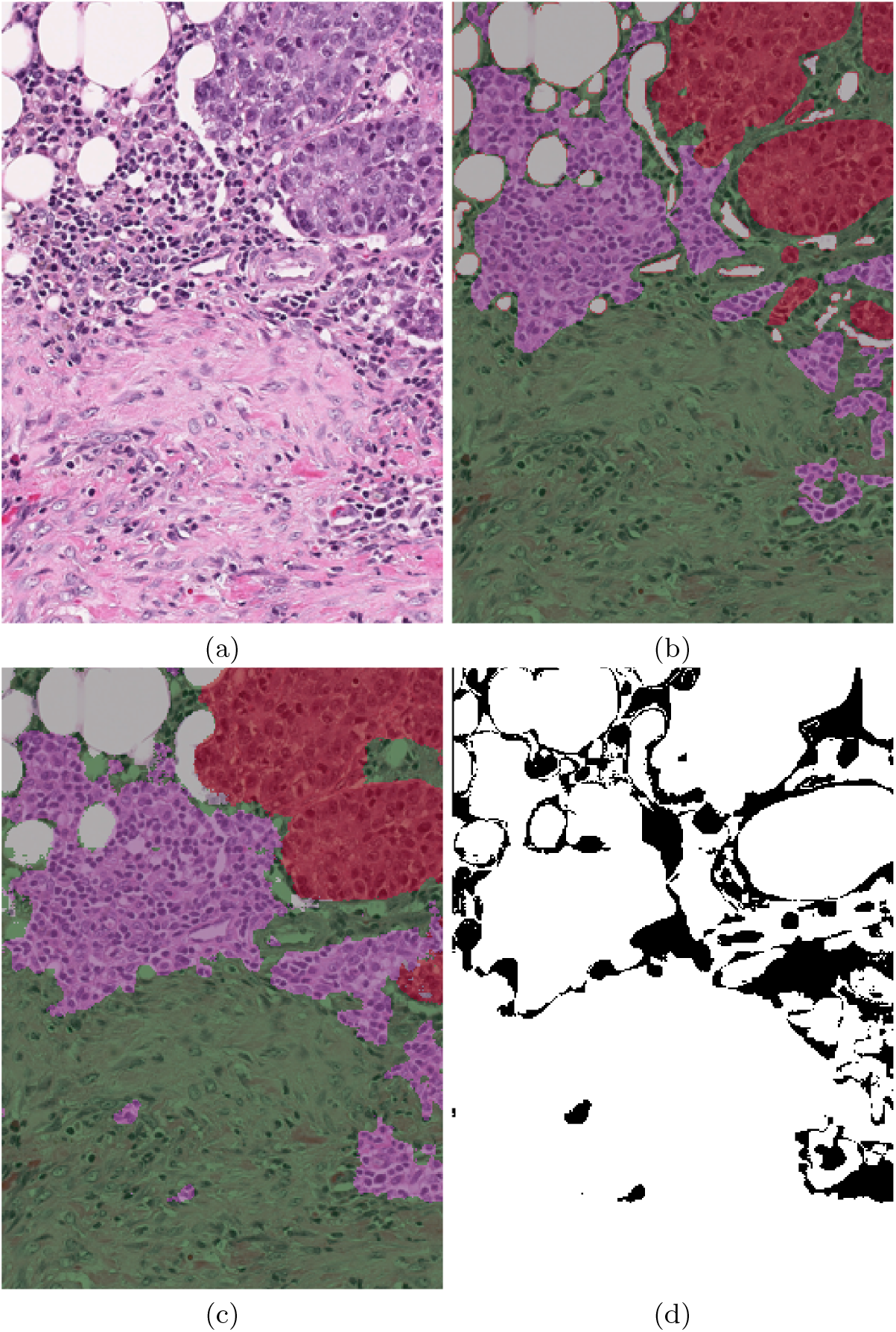
Semantic segmentation into 5 classes (*Tumour, Stroma, Inflammatory, Necrosis, Other*) of the image **TCGA-S3-AA15-DX1** with an overall accuracy of **0.8297**. (a) Original Image (dimensions **2316***×***1864 pixels**). (b) Ground Truth (GT) indicated by shading (Tumour = red, Stroma = green, Inflammatory = purple, Necrosis = blue, Other = gray). (c) Semantic Segmentation. (d) Pixel to pixel comparison of GT and segmentation, white pixels indicate correct class, black indicates incorrect. It should be noted that the errors are mostly in the boundaries between classes.

**Fig. 6.**
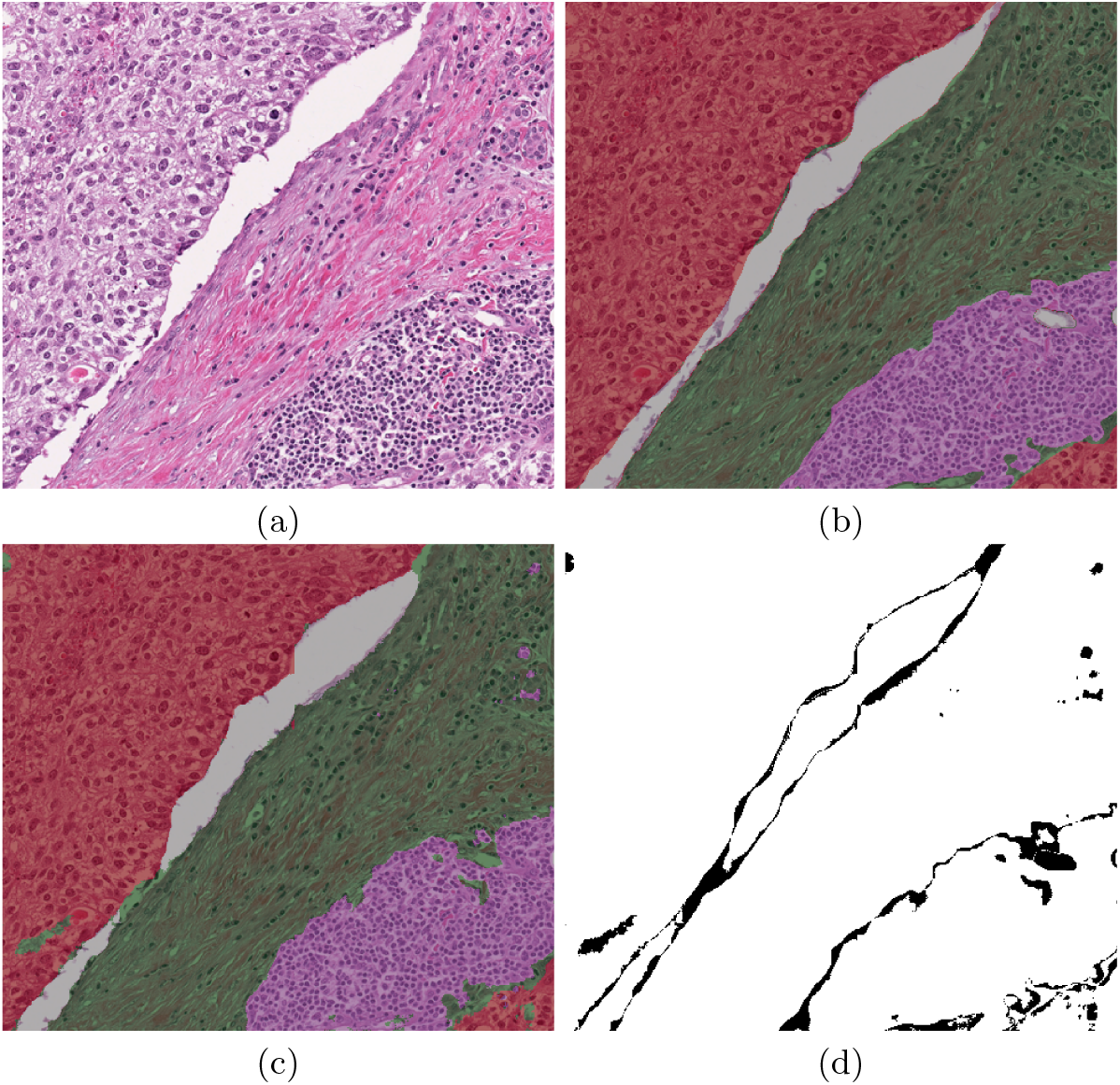
Semantic segmentation into 5 classes (*Tumour, Stroma, Inflammatory, Necrosis, Other*) of the image **TCGA-E2-A1AZ-DX1** with an overall accuracy of **0.9509**. (a) Original Image (dimensions **3047***×***2709 pixels**). (b) Ground Truth (GT) indicated by shading (Tumour = red, Stroma = green, Inflammatory = purple, Necrosis = blue, Other = gray). (c) Semantic Segmentation. (d) Pixel to pixel comparison of GT and segmentation, white pixels indicate correct class, black indicates incorrect. It should be noted that the errors are mostly in the boundaries between classes.

To better understand the overall results, a confusion matrix is presented in Fig. 4. It can be seen that tumour and inflammatory tissue regions outperform the results presented by [1] for Tumour and Inflammatory regions. However in some cases inflammatory is identified as stroma. Inflammatory comprises two regions: lymphocyte and plasma cells which sometimes are confused as benign cells contained within stroma tissue. Also necrosis region can be misinterpreted as stroma region, maybe because sometimes has irregular shape as stroma is most of the time. The remaining tissue sections grouped in *other* have the lowest accuracy in our results, as they are mostly assigned to stroma and tumour. This might be explained as this region includes noise like out of focus, noise artifacts, blood, fat, etc.

## 4 Conclusions

We present a method to perform H&E breast cancer WSI segmentation.In this method segmentation from 4 different tissue regions were evaluated: tumour, stroma, inflammatory, necrosis, and other. Which contrast with methods only dedicated to nuclei and cancer region. Results presented in this paper outperforms results previously reported in two regions: Tumour and Inflammatory. We consider that data provided by BCSS Challenge is a valuable contribution that helps to achieve these results. WSI test images were evaluated with network trained at 50 epochs. It should be noted that whilst we found 50 epochs to report better results than 10 epochs, due to limitations on the computational resources we ran the rest of the experiments with 10 epochs. We can speculate that our results would improve if ran at 50 epochs. Improvements can still be done, first a higher data augmentation can be obtained with an extra overlapping section included during patch extraction. Also, some post processing techniques can be applied to improve results. For example, a method to merge image patches over the sectioned WSI, which motivates to explore in order to rise even more accuracy outcomes.

## Data Availability

All data produced in the present study are available upon reasonable request to the authors

https://github.com/mauOrtRuiz/Breast_Cancer_Sem_Seg

